# Real-World Genetic Characteristics of Candidates for Implantable Cardioverter-Defibrillators with Dilated Cardiomyopathy

**DOI:** 10.1101/2024.03.28.24305043

**Authors:** In-Soo Kim, Yoonjung Kim, Jiwon Seo, Hyemoon Chung, Jung-Woo Son, Sang Jin Ha, Jaemin Shim, Jong-Youn Kim, Se-Joong Rim, Kyung-A Lee, Eui-Young Choi

## Abstract

**Background:** The real-world prevalence of sudden cardiac death (SCD)–related gene mutations and their relationship with implantable cardioverter-defibrillator (ICD) implantation in dilated cardiomyopathy (DCM) has not been clearly investigated.

**Methods:** This study included patients with sporadic DCM with persistent left ventricular ejection fraction (LVEF) ≤ 35% even after 3 months of guideline-directed medical treatment or those with an ICD for secondary prevention and LVEF < 50%. Genetic tests targeting 444 pan-cardiomyopathy genes (including 36 core DCM genes) were performed, along with the collection of clinical and electrophysiological information.

**Results:** A total of 105 patients were enrolled (median age, 65.0 [57.5–75.0] years), and 33 (31%) were women. The average LVEF was 27.4 ± 5.9%, and 46 patients (44%) had ICD or cardiac resynchronization therapy with ICD (34 for primary prevention and 12 for secondary prevention purpose) at the time of enrollment. Fourteen patients (13%) had pathogenic (P) or likely pathogenic (LP) variants (comprising 6 *TTN*, 4 *DSP*, 2 *TNNT2*, 1 *RBM20,* 1 *DSG* and 1 *MYBPC3*, one patients had both *DSP* and *RBM20*), and 59 patients (56%) had P/LP/variant of uncertain significance (VUS) in the 36 core DCM genes. Twenty-eight patients (27%) harbored SCD-related gene variants (14 *DSP*, 11*FLNC*, 9 *RBM20* and 2 *TMEM43*). The prevalence of SCD gene variants tended to be higher in the secondary prevention and non-ICD groups (33.3% for secondary purposes, 11.8% for primary purposes, and 33.8% for non-ICD; p = 0.058).

**Conclusions:** In patients with advanced heart failure and DCM, P/LP-validated DCM variants were not common in sporadic cases. However, when extended to VUS, approximately half of the patients had core DCM-related variants. The prevalence of SCD variants was not lower in the non-ICD group, thus suggesting a potential risk of fatal arrhythmia.

(Trial registration: CRIS KCT0004913)

## Introduction

Dilated cardiomyopathy (DCM) is a disease that accounts for approximately 30% of total heart failure (HF) cases with reduced ejection fraction and is a serious disease with a mortality rate of 20%–30% within 5 years.^1^ However, the exact cause of nonischemic DCM is not well identified, and evidence for genetic contribution is continuously increasing.^2^ Current treatment guidelines around the world state that implantable cardioverter-defibrillators (ICDs) should be inserted for primary prevention purposes in patients with symptomatic DCM with left ventricular ejection fraction (LVEF) < 35%,^3^ but a recently published DANISH trial (researcher-led study) announced that the primary preventive effect of ICD is not different from that of maximum medical treatment in the state of strictly following the guideline-directed medical treatment (GDMT).^4,5^ Complications such as device-related infection or inappropriate defibrillator shock are also issues faced by patients with ICD. After ICD implantation based on LVEF as a primary prevention measure, electrical shock never occurs in 90% of patients, and some patients may receive heart transplants or a left ventricular (LV) assist device (LVAD) within a few years after ICD implantation. However, in the extended follow-up study encompassing 9.6 years, ICD was benefited by a reduction in sudden cardiac death (SCD) in age ≤ 70 years (5% vs. 11%), although events rate was far lower than other previous studies.^5^ It suggests the importance of identifying individuals who are prone to developing early disease onset and progression to advanced HF (i.e., individual risk assessment). Genetic factors are important determinants of early onset and progression to HF in DCM. In addition, specific genetic mutations are known to be related to fatal arrhythmic events.^3^ Therefore, recent cardiomyopathy guidelines emphasize the role of genetic mutations in risk stratification, particularly the risk of sudden arrhythmic death, and provide specific instructions for gene mutation-guided ICD implantation even if LVEF > 35%.^3^ A recent large-scale multicenter study for advanced HF due to DCM showed that the presence of core genetic mutations was related to disease progression to heart transplantation (HT) or LVAD implantation.^6^ However, currently, genetic mutation-guided prophylactic ICD implantation is not popular in real-world practice. Therefore, in this study, we sought to evaluate the prevalence of genetic mutations in ICD candidates for sporadic nonischemic DCM by using core gene and extended pan-cardiomyopathy panels. In real-world practice, we sought to investigate the prevalence of SCD gene mutations in real-world ICD candidates and whether it differed between ICD-implanted patients and nonimplanted patients blinded to genetic information.

## Method

### Study population

Study participants were prospectively enrolled after obtaining agreement for study participation in five tertiary university hospitals (Gangnam Severance Hospital, Wonju Severance Christian Hospital, Korea University Anam Hospital, Kyug Hee University Hospital and Gangneung Asan Hospital). The inclusion criteria for study enrollment were (1) patients who were diagnosed with nonischemic DCM with New York Heart Association (NYHA) functional class II or higher and continuously reduced LVEF of 35% or less than 35% even after 3 months of GDMT after diagnosis regardless of whether primary prevention ICD or cardiac resynchronization therapy (CRT) was performed or (2) patients with nonischemic DCM with LVEF < 50% for more than 3 months after ICD was administered for secondary prevention purposes. Patients with malignant tumors, cerebrovascular accidents, or neuromuscular diseases who were expected to survive within 6 months were excluded from the study. Blood samples for genetic testing were extracted by expert phlebotomists and then transferred to the central core laboratory (Gangnam Severance Hospital Genetic Laboratory) for further analysis. The study protocol was approved by Gangnam Severance and four other centers (IRB No 3-2019-0372, CR319168, 2020AN0454, KHUH2020-03-071 and GNAH2020-01-005), and the clinical data of the participants who agreed to participate in the study were extracted from medical records and entered into a database.

### Clinical data collection

Data on age, sex, history of hypertension and diabetes, HF medication, NYHA functional class, glomerular filtration rate, blood sodium level, and hemoglobin level were collected and entered into a database. A family history of DCM or HF was also obtained. Echocardiographic data such as LV dimension, diastolic functional parameters, left atrial (LA) diameter and volume, and tricuspid regurgitant velocity were collected from echo reports from each center. For the electrocardiogram (ECG) data, presence of atrial fibrillation (AF), QRS duration, bundle branch block (BBB), LV hypertrophy, and number of fragmented QRS in the 12-lead ECG were obtained.

### DNA preparation

Genomic DNA were extracted from ethylenediaminetetraacetic acid-treated whole blood samples via QIAamp DNA Blood Mini kit (Qiagen, Venlo, The Netherlands). Each DNA sample was checked for purity using a NanoDrop 1000 system (Thermo Fisher Scientific, Rockford, IL, USA), and DNA concentration was determined using a Qubit 3.0 fluorometer (Thermo Fisher Scientific). Extracted DNA was stored at -80°C until further use.

### DCM Core 36 gene panel design ^2,7-9^ (Figure 1)

**Figure 1.**
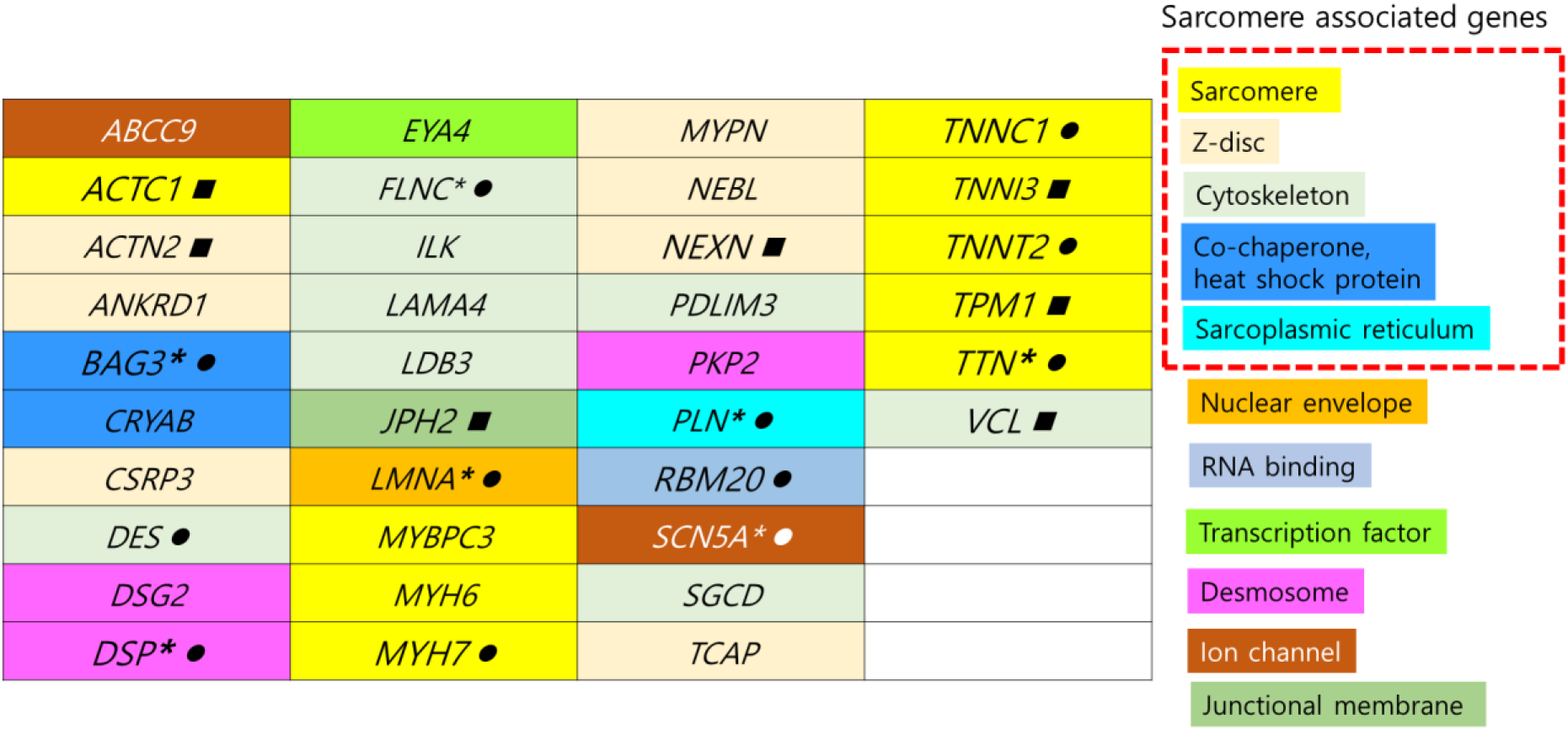
Gene panel for 36 core dilated cardiomyopathy (DCM) variants. Genes with high evidence [12 definitive/strong (*●*) and 7 moderate (*▪*)] according to DCM Gene Curation Expert Panel according to ClinGen.^9^ Those with loss of function as an established DCM-causing mechanism^27^ are noted with an asterisk (*).

To construct the core DCM gene panel, a literature search of the PubMed database was performed for targeted gene selection.^9^

### Pan-cardiomyopathy gene panel ^2,7,8,10,11^ (Supplemental figure 1)

To construct the Pan-cardiomyopathy gene panel, a literature search of the PubMed database was performed for targeted gene selection. Subsequently, a comprehensive Pan-cardiomyopathy gene panel was curated, encompassing 444 genes associated with various cardiomyopathy phenotypes, such as Dilated Cardiomyopathy (DCM), Hypertrophic Cardiomyopathy (HCM), Arrhythmogenic Right Ventricular Cardiomyopathy (ARVC), as well as nuclear DNA genes (MT-nDNA) implicated in mitochondrial cardiomyopathy.

### Library construction and sequencing

For NGS, a library was prepared using the customized Capture probes produced by Celemics, Inc. (Seoul, Korea) to cover coding sequence regions of target genes (Supplemental figure 1). The libraries were prepared using nucleic acid input according to the manufacturer’s instructions. The constructed library was used for sequencing with the Illumina NextSeq 550Dx platform (Illumina, San Diego, CA, USA) with the 2 × 150 base pair paired-end reads.

### Bioinformatics Pipeline for NGS Gene Panel Analysis

Quality control of the sequence reads (FASTQ paired-end reads) were initially performed using FASTQC ^12^. Subsequently, Trimmomatic (version 0.39) ^13^ was employed to trim adapter sequences and eliminate low-quality sequences from the raw sequencing data. The processed reads were aligned to the reference genome (hs37d5) using BWA-MEM (version 0.7.17) ^14^. Picard tools (version 2.27.1) ^15^ were then used to mark duplicated reads and sort BAM files. Variant calling for SNVs and InDels was performed using The Genome Analysis Toolkit (GATK) HaplotypeCaller (version 4.2) ^16^, with VarScan (version 2.4.4) ^17^ serving as complementary method. Bcftools (version 1.10.2) ^18^ was utilized for splitting multi-allelic sites and normalizing variants to produce left-aligned VCF format. Our in-house pipeline, which realigns reads to a target region masked reference genome, was employed to identify variants potentially affected by homologous. ExomeDepth ^19^ was employed for identifying copy number variations.

### Classification of P/LP variants and VUS

To standardize variant interpretation, variants were classified as P, LP or uncertain significance based on American College of Medical Genetics and Genomics standards and guidelines. VCF files were annotated with ANNOVAR^20^ and Variant Effect Predictor. Variants were further filtered with altered allele frequency > 30% and 30x coverage. The variant was classified based on refined American College of Medical Genetics and Genomics (ACMG) standards and specific guidelines for inherited cardiac conditions.^21-24^ The impact of missense change was predicted with REVEL.^25^ REVEL scores >0.644 are more likely to indicate pathogenic variants and scores <0.290 are more likely to indicate benign variants. ^26^ Public databases such as Human Gene Mutation Database professional version, release 2016.1, ClinVar, and Atlas of Cardiac Genetic Variation were used.

### Statistical analysis

The baseline characteristics of the patients are described as mean ± standard deviation for parameters following normal distribution and median (interquartile range) for parameters without normal distribution. The normality of distribution was evaluated using the Shapiro–Wilk test or Kolmogorov– Smirnov test. Differences in clinical features, blood test findings, echocardiography, and electrophysiological findings among ICD implementation groups or genetic mutation groups were analyzed using Student’s t-test or analysis of variance with post hoc analysis. The Mann–Whitney U test and Kruskal–Wallis test were performed for nonparametric variables. Differences, such as the presence or absence of genetic mutations and dichotomous variables, were analyzed using the chi-square test. The analyses were performed using SPSS 27 (IBM Corp), and a two-sided p-value < 0.05 was considered significant.

## Results

### Baseline clinical characteristics

The median age of the patients was 65.0 (57.5–75.0) years, and 33 patients (31%) were women. The average LVEF was 27.4 ± 5.9%, and 46 patients (44%) had ICD or CRT with defibrillator (CRTD) (34 for primary prevention and 12 for secondary prevention purposes). Twenty-nine (28%) patients had persistent AF, and 28 (27%) had BBBs (24 left BBBs and 4 right BBBs). The average LA anteroposterior dimension and E/e’ were 45.0 ± 8.4 mm and 15.2 ± 6.9, respectively. The baseline clinical characteristics of the patients are summarized in **Table 1**.

**Table 1.** Comparisons between the P/LP variant group and others regarding the 36 core DCM genes.

|  | **Total**<br>**(n=105)** | **No P/LP**<br>**variants**<br>**(n=91)** | **Presence of**<br>**P/LP variants in**<br>**Core genes**<br>**(n=14)** | **p** |
| --- | --- | --- | --- | --- |
| **Age, yrs** | 65.0 (57.5 -75.0) | 66 (58 -76) | 63.5 (53-67) | 0.367 |
| **Female, n (%)** | 33 (31) | 27 (30) | 6 (43) | 0.468 |
| **Diabetes, n (%)** | 36 (34) | 30 (33) | 6 (43) | 0.555 |
| **Hypertension, n (%)** | 67 (64) | 56 (62) | 11 (79) | 0.217 |
| **Smoking status,**<br>**quit/current n (%)** | 27/14 (39) | 25/12 (41) | 2/2 (29) | 0.571 |
| **Hb, g/dL** | 13.5±2.1 | 13.6±2.1 | 13.4±2.1 | 0.626 |
| **Na, mEq/mL** | 139 (137-141) | 139 (137-141) | 139.5 (138-141.3) | 0.464 |
| **eGFR, mL/min/1.73m<sup>2</sup>** | 75 (53-91) | 73 (51.3-92.5) | 75 (53-89) | 0.922 |
| **AF, n (%)** | 29 (28) | 26 (29) | 3 (21) | 0.578 |
| **LBBB/RBBB, n (%)** | 24 / 4 (27) | 23/4 (30) | 1/0 (7) | 0.202 |
| **QRS duration, ms** | 106 (97-141) | 110 (98-144) | 101 (91-112) | 0.637 |
| **No of fQRS, n** | 2 (0-5) | 2 (0-6) | 2 (0-3.3) | 0.676 |
| **ICD/CRTD, n (%)** | 33 / 13 (44) | 26 / 13 (43) | 7 / 0 (50) | 0.143 |
| **ICD for 2<sup>nd</sup> prevention** | 12 (11) | 10 (11) | 2 (14) | 0.869 |
| **LVEF, %** | 27.4±5.9 | 27.5±6.2 | 27.3±5.1 | 0.869 |
| **LVEDD, mm** | 61.4±7.9 | 61.3±8.2 | 61.5±7.1 | 0.890 |
| **LV mass index, g/m<sup>2</sup>** | 139.8±39.7 | 138.2±42.1 | 144.0±36.4 | 0.524 |
| **s', cm/s** | 4 (3-5) | 4 (3-5) | 3.5 (2.8-4.2) | 0.559 |
| **E, cm/s** | 71.0 (52.5-97.8) | 71 (54-96.3) | 57.5 (34.4-96.3) | 0.257 |
| **e', cm/s** | 5 (4-6) | 5 (4-6) | 4 (2.9-5) | 0.823 |
| **E/e'** | 14.0 (10.8-19.2) | 13.5 (10.4-19.0) | 15.9 (10.8-21.0) | 0.050 |
| **Functional MR, n** | 2 (1-2) | 2 (1-2) | 2 (1-2) | 0.994 |
| **LA dimension, mm** | 45.0±8.4 | 45.6±7.6 | 44.5±9.0 | 0.606 |
| **LAVI, mL/m<sup>2</sup>** | 48.1 (34.7-61.2) | 48.0 (35.0-59.1) | 49.5 (27.9-67.8) | 0.660 |
| **TRV max, m/s** | 2.3 (2.1-2.7) | 2.3 (2.1-2.6) | 2.3 (2.1-2.8) | 0.607 |
| **ARNI, n (%)** | 80 (76) | 68 (75) | 12 (86) | 0.369 |
| **ARB/ACEi, n (%)** | 24 (23) | 23 (25) | 1(7) | 0.133 |
| **Beta blockers, n (%)** | 91 (87) | 78 (86) | 13 (93) | 0.464 |
| **MRA, n (%)** | 58 (55) | 51 (56) | 7 (50) | 0.672 |

eGFR, estimated glomerular filtration rate; LBBB, left bundle branch block; RBBB, right bundle branch block; fQRS, fragmented QRS; ICD, implantable cardioverter–defibrillator; CRTD, cardiac resynchronization therapy with defibrillator; LA, left atrial; LAVI, LA volume index; TRV, tricuspid regurgitant velocity; ARB, angiotensin II receptor blocker; ARNI, angiotensin II receptor and neprilysin inhibitor; MRA, mineralocorticoid receptor antagonist; functional MR, functional mitral regurgitation (0 = none, 1 = trivial, 2 = mild, 3 = moderate, 4 = severe).

**Table 2.** Comparison between the 36 core DCM variants group and the no variants group up to VUS.

|  | **No core variants**<br>(n = 46) | **36 core DCM variants**<br>(n = 59) | **p** |
| --- | --- | --- | --- |
| **Age, yrs** | 68 (58.0 -78.0) | 64.5 (56.3-74.3) | 0.391 |
| **Female, n (%)** | 14 (30) | 19 (32) | 0.846 |
| **Diabetes, n (%)** | 17 (37) | 19 (32) | 0.611 |
| **Hypertension, n (%)** | 27 (59) | 40 (68) | 0.244 |
| **Smoking status**<br>quit/current, (%) | 10/4 (30) | 17/10 (46) | 0.306 |
| **Hb, g/dL** | 13.5±2.3 | 13.5±1.9 | 0.968 |
| **Na, mEq/mL** | 139 (136-141) | 139 (137-141) | 0.730 |
| **eGFR, mL/min/1.73m<sup>2</sup>** | 71.1 (49.1-92.6) | 79.6 (55.4-90.6) | 0.504 |
| **AF, n (%)** | 15 (33) | 14 (24) | 0.313 |
| **LBBB/RBBB, n (%)** | 11 / 3 (30) | 13/1 (24) | 0.411 |
| **QRS duration, ms** | 106 (92-150) | 110 (100-134) | 0.660 |
| **No of fQRS, n** | 1 (0-5.3) | 2 (0-5) | 0.449 |
| **ICD/CRTD, n (%)** | 14 / 8 (48) | 19 / 5 (40) | 0.383 |
| **ICD for 2<sup>nd</sup> prevention** | 5 (11) | 7 (12) | 0.675 |
| **LVEF, %** | 28.3±6.7 | 26.7±5.2 | 0.170 |
| **LVEDD, mm** | 61.6±8.7 | 61.2±7.3 | 0.828 |
| **LV mass index, g/m<sup>2</sup>** | 140.4±44.0 | 139.2±38.0 | 0.878 |
| **s', cm/s** | 4 (3-5) | 4 (3-5) | 0.777 |
| **E, cm/s** | 69 (50.5-84.8) | 69 (51.8-99.3) | 0.675 |
| **e', cm/s** | 6 (4-6.8) | 5 (4-6) | 0.121 |
| **E/e'** | 12.8 (9.8-18.5) | 14.0 (11.2-20.5) | 0.157 |
| **Functional MR, n** | 2 (1-2.3) | 2 (1-2) | 0.581 |
| **LA dimension, mm** | 45.4±7.5 | 44.6±9.1 | 0.647 |
| **LAVI, mL/m<sup>2</sup>** | 51.2 (35.0-59.1) | 46.5 (33.6-63.8) | 0.718 |
| **TRV max, m/s** | 2.3 (2.2-2.7) | 2.3 (2.0-2.7) | 0.711 |
| **ARNI, n (%)** | 32 (70) | 48 (81) | 0.159 |
| **ARB/ACEi, n (%)** | 12 (26) | 12 (20) | 0.486 |
| **Beta blockers, n (%)** | 37 (80) | 54 (92) | 0.097 |
| **MRA, n (%)** | 25 (54) | 33 (56) | 0.871 |

See abbreviations in table 1.

### Genetic test results in targeted 36 core DCM genes and 444 pan-cardiomyopathy genes

Fourteen (13%) patients had P or LP variants (six *TTN*, four *DSP*, two *TNNT2,* one *RBM20,* one *DSG2, and* one *MYBPC3,* one patient had both *DSP*, and *RBM20*), and 59 (56%) had P/LP/variant of uncertain significance (VUS) in the 36 core DCM panel. The three most common variants up to the VUS were *TTN*, *DSP* and *FLNC*. The variant coding proteins comprised 36% sarcomere; 20% desmosome; 18% cytoskeleton; 11% Z-disk; 8% ribonucleic acid binding; 3% junctional membrane and 4% ion channels, co-chaperones, or transcription factors **(Figure 2)**. When extended to 444 pan-cardiomyopathy-associated gene panels, 34 (32%) patients had P/LP variants, and 102 (97%) had P/LP/VUS. The P/LP variants detected in the pan-cardiomyopathy panel comprised two *HFE*, two *ACADS*, one *HPS1*, one *GNE*, one *OPA1*, three *GAA*, two *STAR*, one *TREX1*, one *NEK8*, one *FKRP*, one *GLB1*, one *FANCI*, one *ANO5*, one *TMEM126A*, one *GNPTAB*, two *GNE*, one *ALMS1*, one *NUP107*, one *IFIH1*, and one *WFS1* variant.

**Figure 2.**
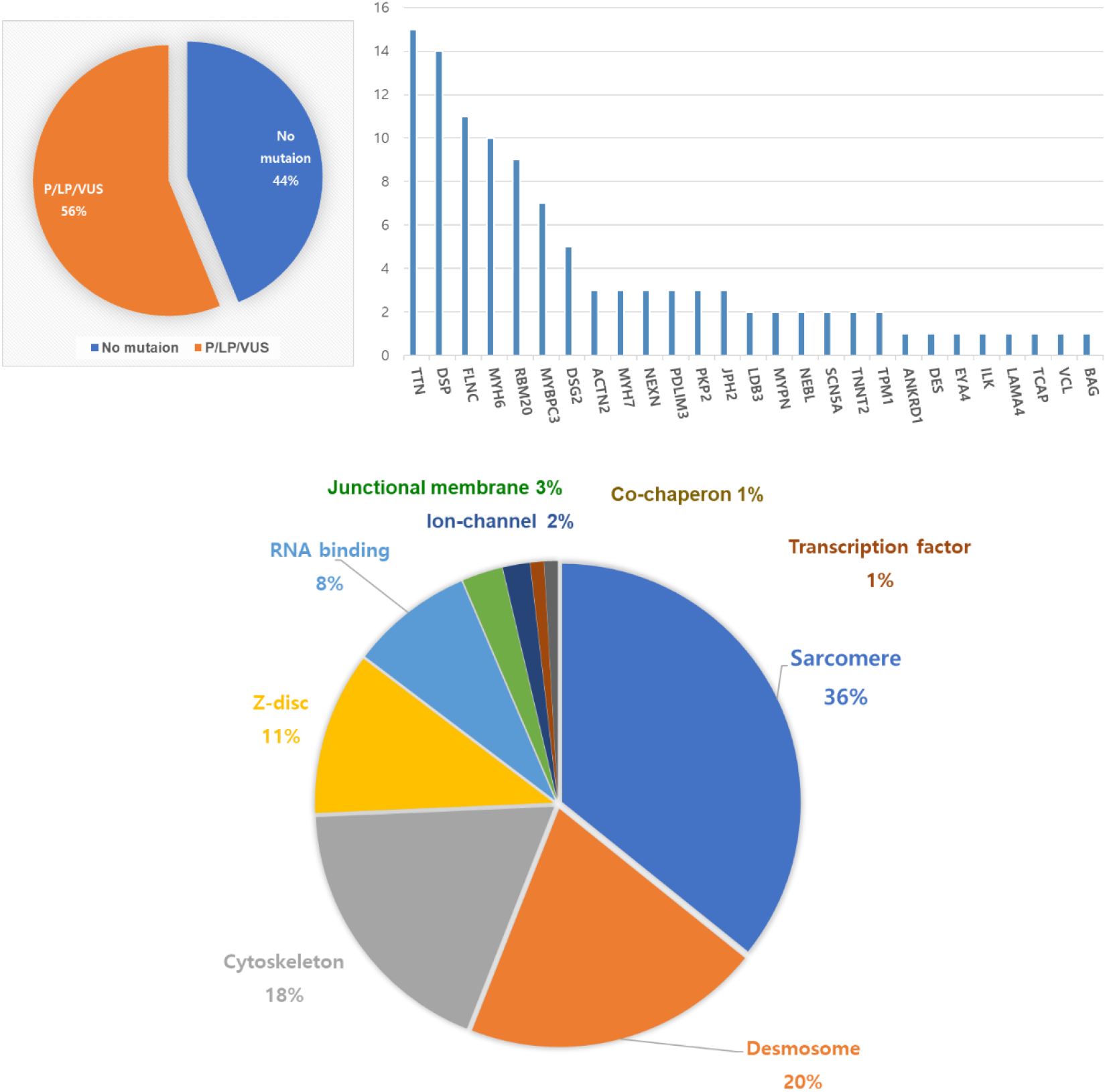
Distribution and prevalence of P/LP/VUS in the 36 core DCM-associated genes.

### Clinical, echocardiography, and electrophysiological findings according to genetic mutation

When comparing patients with P/LP variants (in the 36 core and 444 pan-cardiomyopathy panels) and others for clinical, echocardiographic, and electrophysiological findings, patients with P/LP variants were younger, were predominantly females, and had reduced diastolic and systolic functions; however, statistical significance was not reached. When extended to VUS, patients with LP/P or VUS showed similar trends, but statistical significance was not reached **(Tables 1–3)**. Patients with SCD-related variants according to the European Society of Cardiology cardiomyopathy guidelines were 28 (27%) up to VUS. They comprised 14 *DSP*, 11 *FLNC*, 9 *RBM20*, and 2 *TMEM43* variants (three patients had more than one SCD variant); however, no *LMNA* variants were detected in this population. The prevalence of SCD gene variants tended to be higher in the secondary prevention ICD group than in the primary prevention ICD group (33.3% for secondary purposes, 11.8% for primary purposes, and 33.8% for no ICD; p = 0.058) **(Figure 3)**. An interesting finding was that the prevalence of SCD variants was not lower in the non-ICD group than in the secondary prevention ICD group, thus suggesting exposure to the risk of SCD. However, there were no significant differences in the prevalence of AF and BBB between the SCD variant groups and the other groups. Interestingly, the prevalence of ICD/CRTD implantation was lower in patients with SCD variants (30% in the SCD variants group and 49% in the other groups, p = 0.162) **(Table 4)**. The reason for non-ICD implantation was the patient denial or doctors’ decision to wait, thus suggesting that it was not based on scientific background. When comparing the sarcomere gene mutation-positive group (n = 29, 28%) with the other groups, no significant differences were observed in the echocardiography and clinical characteristics **(Table 5)**.

**Figure 3.**
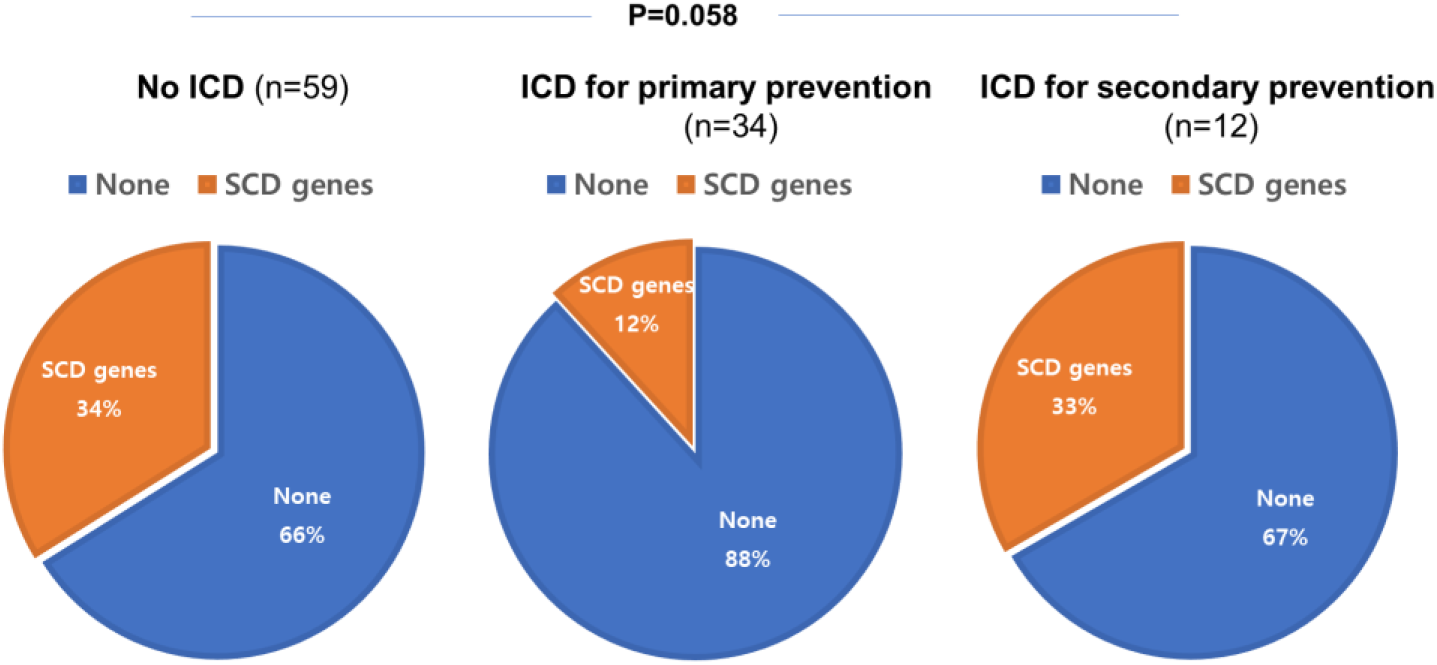
Prevalence of sudden cardiac death (SCD) gene variants according to implantable cardioverter defibrillator (ICD) implantation.

**Table 3.** Comparison between P/LP variants and others in the 444 pan-cardiomyopathy gene panel.

|  | **No P/LP variants**<br>**(n=71)** | **Presence of**<br>**P/LP variants**<br>**(n=34)** | **p** |
| --- | --- | --- | --- |
| **Age, yrs** | 67 (59 -76) | 63.5 (53-71.3) | 0.367 |
| **Female, n (%)** | 17 (24) | 16 (47) | 0.017 |
| **Diabetes, n (%)** | 23 (32) | 13 (38) | 0.555 |
| **Hypertension, n (%)** | 30 (42) | 8 (24) | 0.062 |
| **Smoking status,** | 24/8 (45) | 3/6 (27) | 0.023 |
| **Hb, g/dL** | 13.6±2.1 | 13.4±2.1 | 0.626 |
| **Na, mEq/mL** | 139 (137-141) | 139 (138-140) | 0.464 |
| **eGFR, mL/min/1.73m<sup>2</sup>** | 76.5 (53.1-91.8) | 75 (53-89) | 0.922 |
| **AF, n (%)** | 21 (30) | 8 (24) | 0.517 |
| **LBBB/RBBB, n (%)** | 17/2 (27) | 7/2 (27) | 0.712 |
| **QRS duration, ms** | 110 (96-144) | 105 (93.5-139) | 0.637 |
| **No of fQRS, n** | 2 (0-6) | 2 (0-4) | 0.676 |
| **ICD/CRTD, n (%)** | 21 / 11 (45) | 12 / 2 (40) | 0.365 |
| **ICD for 2<sup>nd</sup> prevention** | 6 (9) | 6 (18) | 0.225 |
| **LVEF, %** | 27.5±6.2 | 27.3±5.1 | 0.869 |
| **LVEDD, mm** | 61.3±8.2 | 61.5±7.1 | 0.890 |
| **LV mass index, g/m<sup>2</sup>** | 138.2±42.1 | 144.0±36.4 | 0.524 |
| **s', cm/s** | 4 (3-5) | 4 (4-5) | 0.559 |
| **E, cm/s** | 67 (49-87) | 71 (56-110) | 0.257 |
| **e', cm/s** | 5 (4-6) | 5 (4-6) | 0.823 |
| **E/e'** | 13.1 (10.0-17.5) | 16.2 (13.1-21.7) | 0.050 |
| **Functional MR, n** | 2 (1-2) | 2 (1-2) | 0.994 |
| **LA dimension, mm** | 45.6±7.6 | 44.5±9.0 | 0.606 |
| **LAVI, mL/m<sup>2</sup>** | 48.1 (34.2-59.3) | 48.6 (37.7-64.9) | 0.660 |
| **TRV max, m/s** | 2.3 (2.1-2.6) | 2.4 (2.1-2.8) | 0.607 |
| **ARNI, n (%)** | 52 (73) | 28 (82) | 0.305 |
| **ARB/ACEi, n (%)** | 19 (27) | 5 (15) | 0.169 |
| **Beta blockers, n (%)** | 64 (90) | 27 (79) | 0.130 |
| **MRA, n (%)** | 40 (56) | 18 (53) | 0.743 |

See abbreviations in table 1.

**Table 4.**
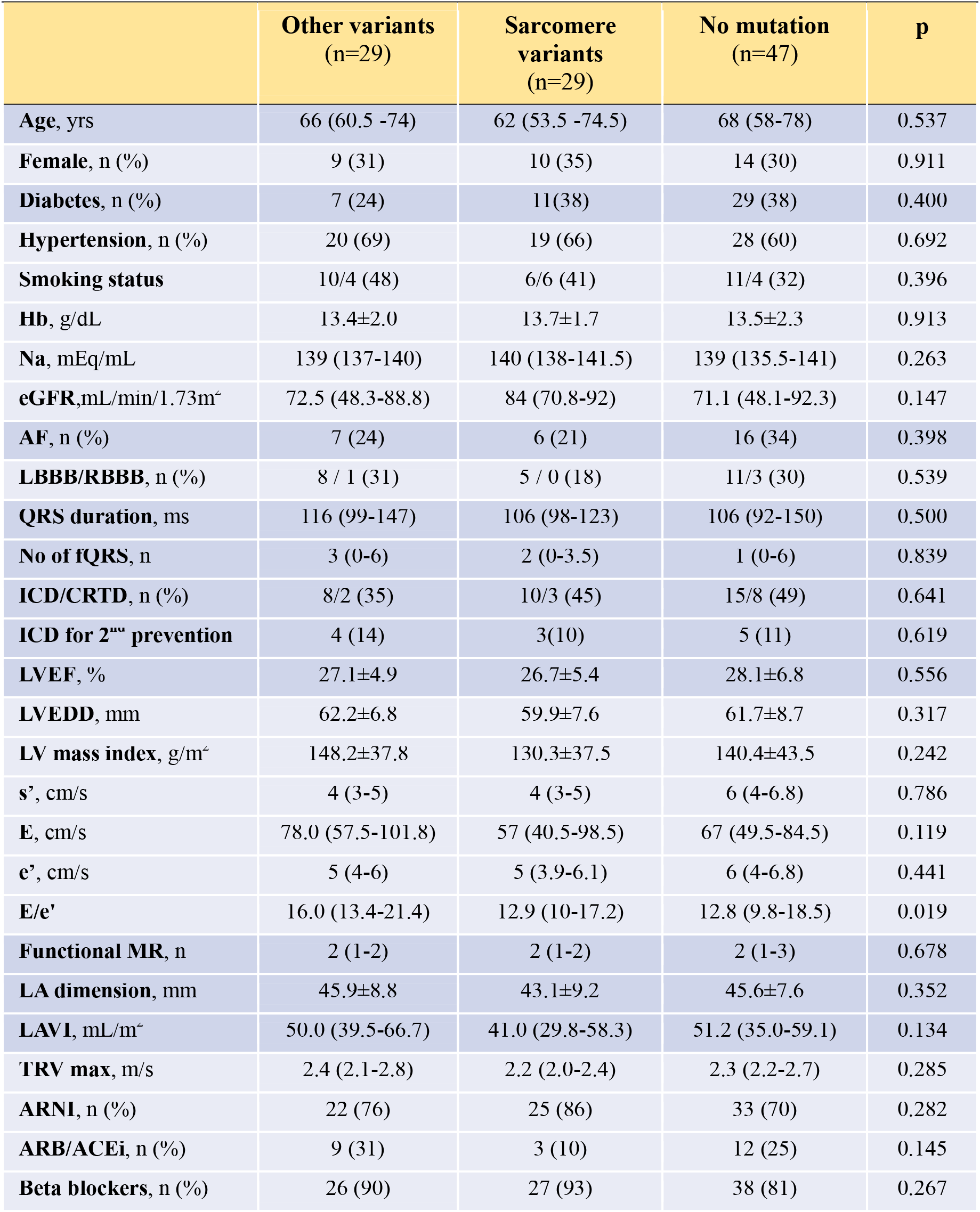
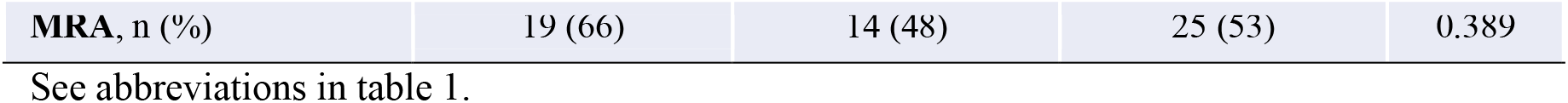
Comparisons between sarcomere-related gene mutation and others for clinical characteristics.

|  | **Other variants**<br>**(n=29)** | **Sarcomere**<br>**variants**<br>**(n=29)** | **No mutation**<br>**(n=47)** | **p** |
| --- | --- | --- | --- | --- |
| **Age, yrs** | 66 (60.5 -74) | 62 (53.5 -74.5) | 68 (58-78) | 0.537 |
| **Female, n (%)** | 9 (31) | 10 (35) | 14 (30) | 0.911 |
| **Diabetes, n (%)** | 7 (24) | 11(38) | 29 (38) | 0.400 |
| **Hypertension, n (%)** | 20 (69) | 19 (66) | 28 (60) | 0.692 |
| **Smoking status** | 10/4 (48) | 6/6 (41) | 11/4 (32) | 0.396 |
| **Hb, g/dL** | 13.4±2.0 | 13.7±1.7 | 13.5±2.3 | 0.913 |
| **Na, mEq/mL** | 139 (137-140) | 140 (138-141.5) | 139 (135.5-141) | 0.263 |
| **eGFR,mL/min/1.73m<sup>2</sup>** | 72.5 (48.3-88.8) | 84 (70.8-92) | 71.1 (48.1-92.3) | 0.147 |
| **AF, n (%)** | 7 (24) | 6 (21) | 16 (34) | 0.398 |
| **LBBB/RBBB, n (%)** | 8 / 1 (31) | 5 / 0 (18) | 11/3 (30) | 0.539 |
| **QRS duration, ms** | 116 (99-147) | 106 (98-123) | 106 (92-150) | 0.500 |
| **No of fQRS, n** | 3 (0-6) | 2 (0-3.5) | 1 (0-6) | 0.839 |
| **ICD/CRTD, n (%)** | 8/2 (35) | 10/3 (45) | 15/8 (49) | 0.641 |
| **ICD for 2<sup>nd</sup> prevention** | 4 (14) | 3(10) | 5 (11) | 0.619 |
| **LVEF, %** | 27.1±4.9 | 26.7±5.4 | 28.1±6.8 | 0.556 |
| **LVEDD, mm** | 62.2±6.8 | 59.9±7.6 | 61.7±8.7 | 0.317 |
| **LV mass index, g/m<sup>2</sup>** | 148.2±37.8 | 130.3±37.5 | 140.4±43.5 | 0.242 |
| **s', cm/s** | 4 (3-5) | 4 (3-5) | 6 (4-6.8) | 0.786 |
| **E, cm/s** | 78.0 (57.5-101.8) | 57 (40.5-98.5) | 67 (49.5-84.5) | 0.119 |
| **e', cm/s** | 5 (4-6) | 5 (3.9-6.1) | 6 (4-6.8) | 0.441 |
| **E/e'** | 16.0 (13.4-21.4) | 12.9 (10-17.2) | 12.8 (9.8-18.5) | 0.019 |
| **Functional MR, n** | 2 (1-2) | 2 (1-2) | 2 (1-3) | 0.678 |
| **LA dimension, mm** | 45.9±8.8 | 43.1±9.2 | 45.6±7.6 | 0.352 |
| **LAVI, mL/m<sup>2</sup>** | 50.0 (39.5-66.7) | 41.0 (29.8-58.3) | 51.2 (35.0-59.1) | 0.134 |
| **TRV max, m/s** | 2.4 (2.1-2.8) | 2.2 (2.0-2.4) | 2.3 (2.2-2.7) | 0.285 |
| **ARNI, n (%)** | 22 (76) | 25 (86) | 33 (70) | 0.282 |
| **ARB/ACEi, n (%)** | 9 (31) | 3 (10) | 12 (25) | 0.145 |
| **Beta blockers, n (%)** | 26 (90) | 27 (93) | 38 (81) | 0.267 |
| **MRA, n (%)** | 19 (66) | 14 (48) | 25 (53) | 0.389 |

See abbreviations in table 1.

**Table 5.** Comparison between patients with SCD gene variants and others.

|  | **No SCD gene variants (n=77)** | **SCD gene variants (n=28)** | **p** |
| --- | --- | --- | --- |
| **Age, yrs** | 64 (57 -75.5) | 65.5 (61-75) | 0.897 |
| **Female, n (%)** | 21 (27) | 12 (43) | 0.128 |
| **Diabetes, n (%)** | 18 (23) | 9 (32) | 0.457 |
| **Hypertension, n (%)** | 49 (64) | 18 (64) | 0.951 |
| **Smoking status,** | 18/12 (39) | 9/2 (39) | 0.425 |
| **Hb, g/dL** | 13.6±2.1 | 13.4±2.1 | 0.626 |
| **Na, mEq/mL** | 139 (137-141) | 139 (138-140) | 0.464 |
| **eGFR,mL/min/1.73m<sup>2</sup>** | 76.5 (53.1-91.8) | 75 (53-89) | 0.922 |
| **AF, n (%)** | 23 (30) | 6 (21) | 0.392 |
| **LBBB/RBBB, n (%)** | 18/3 (27) | 6/2 (24) | 0.449 |
| **QRS duration, ms** | 110 (96-144) | 105 (93.5-139) | 0.637 |
| **No of fQRS, n** | 2 (0-6) | 2 (0-4) | 0.676 |
| **ICD/CRTD, n (%)** | 27 / 11 (49) | 6 / 2 (29) | 0.162 |
| **ICD for 2<sup>nd</sup> prevention** | 8 (10) | 4 (14) | 0.058 |
| **LVEF, %** | 27.5±6.2 | 27.3±5.1 | 0.869 |
| **LVEDD, mm** | 61.3±8.2 | 61.5±7.1 | 0.890 |
| **LV mass index, g/m<sup>2</sup>** | 138.2±42.1 | 144.0±36.4 | 0.524 |
| **s', cm/s** | 4 (3-5) | 4 (4-5) | 0.559 |
| **E, cm/s** | 67 (49-87) | 71 (56-110) | 0.257 |
| **e', cm/s** | 5 (4-6) | 5 (4-6) | 0.823 |
| **E/e'** | 13.1 (10.0-17.5) | 16.2 (13.1-21.7) | 0.050 |
| **Functional MR, n** | 2 (1-2) | 2 (1-2) | 0.994 |
| **LA dimension, mm** | 45.6±7.6 | 44.5±9.0 | 0.606 |
| **LAVI, mL/m<sup>2</sup>** | 48.1 (34.2-59.3) | 48.6 (37.7-64.9) | 0.660 |
| **TRV max, m/s** | 2.3 (2.1-2.6) | 2.4 (2.1-2.8) | 0.607 |
| **ARNI, n (%)** | 58 (75) | 22 (79) | 0.195 |
| **ARB/ACEi, n (%)** | 16 (21) | 8 (29) | 0.400 |
| **Beta blockers, n (%)** | 66 (86) | 25 (89) | 0.634 |
| MRA, n (%) | 40 (52) | 18 (64) | 0.261 |

See abbreviations in Table 1.

## Discussion

We found that the prevalence of P/LP variants was not as high as 14% in the 36-core DCM panel and 32% in the 444 pan-cardiomyopathy panels in ICD candidates with sporadic DCM. However, when extended to VUS, the prevalence of genetic mutations significantly increased to 56% in the core DCM panel and 97% in the pan-cardiomyopathy panel, thus suggesting a potential genetic background. Patients tended to be younger, were predominantly female, and had poorer systolic and diastolic function. In addition, the prevalence of SCD-related variants (*LMNA*, *FLNC*, *DSP*, *PLN*, *TMEM43*, and *RBM20*) was higher in patients with ICD for secondary prevention than in those with ICD for primary prevention. However, the prevalence was not lower in the non-ICD group (34%) than in the secondary prevention group, thus suggesting a potential risk of fatal arrhythmia.

### Prevalence and distribution of genetic variants in ICD candidates of DCM

Even after the exclusion of the ischemic and reversible causes of systolic LV dysfunction, the prevalence of P or LP variants in the core genes was low in patients with sporadic DCM. Although we further extended the panel to 444 genes, the prevalence of any P/LP variant was not as high as 32%. However, when we included VUS, the prevalence of the core DCM genes was 56%. Although the role of the genes included in the extended 444 pan-cardiomyopathy panel was not clear in DCM, almost all patients (97 %) had VUS. This suggests that a large proportion of patients had genetic mutations in DCM with persistent LV systolic dysfunction or a history of fatal arrhythmic events. Although the expert committee recommended limiting the genetic panel size because of the ambiguity of pathogenicity and concerns about family genetic counseling^9^, the contribution of various genes to the development or phenotypes of DCM has not been widely investigated compared with hypertrophic cardiomyopathy. The current study detected not only sarcomere-related mutations but also other mutations that encode various small organelles. Although the most common mutation was *TTN*, which is a representative variant of DCM, other gene mutations related to the cytoskeleton, sarcoplasmic reticulum, nuclear envelope, RNA-binding site, desmosome, and ion channels were also detected.^27^ This suggests that multiple pathways are involved in the development of DCM and progression to advanced HF. Furthermore, this indicates the complexity of gene-based targeted therapy in DCM. In the current study, the main pathogenic variant was *TTN*, which has been shown to be relatively benign,^27^ and the other important pathogenic gene variant was *DSP*. Although the *DSP* variants are known to be related to arrhythmogenic right ventricular cardiomyopathy (ARVC),^28^ studies have shown common pathogenicity in ARVC and DCM, thus indicating that they share a genetic contribution.^3,9^ Therefore, in the current study, the interpretation of *DSP* followed the ARVC pathogenicity criteria. We also identified some roles of *DSP* in the development and progression of DCM.

### Clinical, echocardiographic, and electrophysiological characteristics according to mutation

In our study, patients with core mutations up to VUS were younger, thus suggesting early disease onset and early progression to advanced HF. They tended to have lower systolic and diastolic functions without statistical significance owing to the small population size. This finding is compatible with that of a recent study by Hofmeyer et al. about the worse prognosis and higher prevalence of ICD/LVAD/HT in mutation-positive patients.^6^ However, in this relatively homogenous group of patients (97% had LVEF < 35% even after 3 months of GDMT), specific genetic mutations were not related to LV systolic and diastolic dysfunction, BBB, and AF. In the extended pan-cardiomyopathy panel, patients with P/LP variants were predominantly female, had a lower rate of history of hypertension, and were younger but not significantly, thus suggesting a meaningful genetic contribution. We also grouped the patients according to sarcomere mutation for LV systolic and diastolic functions and according to the presence of SCD-related genes for electrophysiological characteristics such as AF, BBB, or number of fragmented QRS in a 12-lead ECG. However, this failed to reach a statistically significant difference among groups. This suggests that specific mutations do not contribute to myocardial function or electrophysiological properties. Rather, the burden of genetic mutations contributes to early disease progression and eventually results in LV dysfunction and electrophysiological instability.

### SCD genes and real-world practice in ICD candidates of DCM

A recent cardiomyopathy guideline suggested seven SCD gene mutations: *LMNA*, *FLNC* truncating, *PLN*, *DSP*, *RBM20*, and *TMEM43* mutations.^3^ Although gene mutations were not common, *DSP*, *FLNC truncating*, *RBM20*, and *TMEM43* VUS were detected in several patients in the current study. However, in our study, the *LMNA* variant, which is known to be associated with the worst prognosis^29^ was not detected. This might be due to the nature of the relatively low admission of sick patients in outpatient clinics or general wards. In our study population, the ICD or CRTD implantation rate of the patients was only 53% even for the Ia or IIa recommendation by current guidelines.^3^ It might be due to the effects of the DANISH trial^4^ or recent improvement of GDMT such as sacubitril/valsartan or sodium-glucose cotransporter-2 inhibitors; therefore, structural and functional reverse remodeling can be expected after 3 months or patient’s denial. We found that the prevalence of SCD gene mutations tended to be higher in patients with ICD for secondary prevention than in those with ICD for primary prevention. Interestingly, patients without ICD had a higher prevalence of SCD gene mutations in real practice in a blinded situation, thus suggesting exposure to the risk of fatal arrhythmias. Regarding ICD implantation in patients with LVEF < 35%, a genetic test would reinforce to patients and doctors the need for ICD implantation.

### Study limitation

First, the study population was relatively homogenous because almost all patients had an LVEF of 35% or <35% despite the three-month GDMT. Therefore, the differences in phenotypic expression based on genetic mutations are very narrow. Future long-term follow-up studies are needed to determine the clinical outcomes. Second, in patients with sporadic DCM, the prevalence of pathogenic mutations is low; therefore, more patients are needed to identify genetic mutation-related phenotypic differences.

## Conclusion

In patients with advanced HF and DCM, P/LP-validated DCM variants were not common in sporadic cases; however, when extended to VUS, more than half of the patients had DCM-related gene variants. In the advanced HF with DCM group, patients with any related variants had early disease onset. The prevalence of SCD-related variants tended to be higher in patients with ICDs for secondary prevention than in those with ICDs for primary prevention. However, the prevalence was not lower in the non-ICD group (34%), thus suggesting a potential risk of fatal arrhythmia. This suggests that genetic testing for SCD genes is required before a final decision is made for ICD implantation for primary prevention.

## Data Availability

The datasets used and/or analyzed in the current study are available from the corresponding author upon reasonable request.

## Nonstandard Abbreviations and Acronyms

DCM: dilated cardiomyopathy
GDMT: guideline-directed medical treatment
SCD: sudden cardiac death
NYHA: New York Heart Association
CRT: cardiac resynchronization therapy
P: pathogenic
LP: likely pathogenic
MAF: minor allele frequency
CRTD: CRT with defibrillator
BBB: bundle branch block
VUS: variant of uncertain significance
ARVC: arrhythmogenic right ventricular cardiomyopathy

## Acknowledgments

None

## Sources of Funding

This work was supported by the Basic Science Research Program through the National Research Foundation of Korea (NRF) funded by the Ministry of Education (2019R1F1A1045911) and Research Fund of Korean Society of Circulation, (202003-04).

## Disclosures

None

## Supplemental Figure

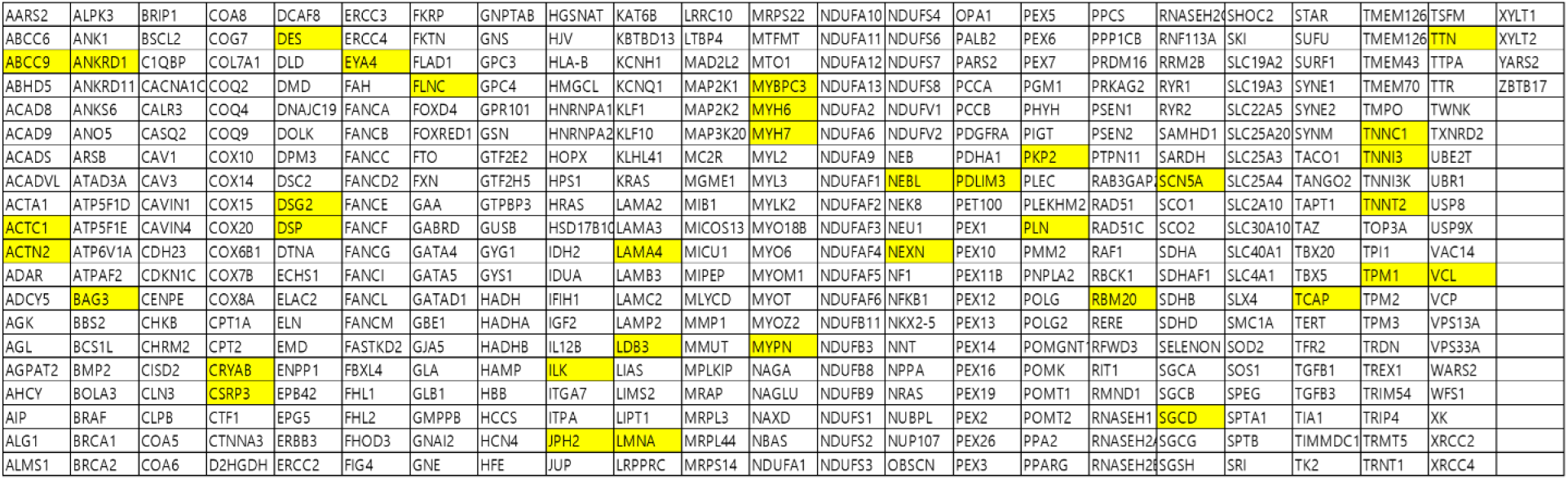

The 444 pan-cardiomyopathy gene panel. Yellow box indicates 36 core DCM associated genes.

## Notes

### Competing Interest Statement

The authors have declared no competing interest.

### Clinical Trial

CRIS KCT0004913

### Author Declarations

The study protocol was approved by Gangnam Severance and four other centers (IRB 17 No 3-2019-0372, CR319168, 2020AN0454, KHUH2020-03-071 and GNAH2020-01-005)

## References

1. Schultheiss H-P, Fairweather D, Caforio ALP, Escher F, Hershberger RE, Lipshultz SE, Liu PP, Matsumori A, Mazzanti A, McMurray J, et al. Dilated cardiomyopathy. Nature Reviews Disease Primers. 2019;5:32. doi: 10.1038/s41572-019-0084-1

2. McNally EM, Mestroni L. Dilated Cardiomyopathy: Genetic Determinants and Mechanisms. Circ Res. 2017;121:731–748. doi: 10.1161/circresaha.116.309396

3. Arbelo E, Protonotarios A, Gimeno JR, Arbustini E, Barriales-Villa R, Basso C, Bezzina CR, Biagini E, Blom NA, de Boer RA, et al. 2023 ESC Guidelines for the management of cardiomyopathies: Developed by the task force on the management of cardiomyopathies of the European Society of Cardiology (ESC). European Heart Journal. 2023;44:3503–3626. doi: 10.1093/eurheartj/ehad194

4. Køber L, Thune JJ, Nielsen JC, Haarbo J, Videbæk L, Korup E, Jensen G, Hildebrandt P, Steffensen FH, Bruun NE, et al. Defibrillator Implantation in Patients with Nonischemic Systolic Heart Failure. New England Journal of Medicine. 2016;375:1221–1230. doi: 10.1056/NEJMoa1608029

5. Yafasova A, Butt JH, Elming MB, Nielsen JC, Haarbo J, Videbæk L, Olesen LL, Steffensen FH, Bruun NE, Eiskjær H, et al. Long-Term Follow-Up of DANISH (The Danish Study to Assess the Efficacy of ICDs in Patients With Nonischemic Systolic Heart Failure on Mortality). Circulation. 2022;145:427–436. doi: doi:10.1161/CIRCULATIONAHA.121.056072

6. Hofmeyer M, Haas GJ, Jordan E, Cao J, Kransdorf E, Ewald GA, Morris AA, Owens A, Lowes B, Stoller D, et al. Rare Variant Genetics and Dilated Cardiomyopathy Severity: The DCM Precision Medicine Study. Circulation. 2023;148:872–881. doi: 10.1161/circulationaha.123.064847

7. Fu Y, Eisen HJ. Genetics of Dilated Cardiomyopathy. Current Cardiology Reports. 2018;20:121. doi: 10.1007/s11886-018-1061-0

8. Tayal U, Prasad S, Cook SA. Genetics and genomics of dilated cardiomyopathy and systolic heart failure. Genome Med. 2017;9:20. doi: 10.1186/s13073-017-0410-8

9. Jordan E, Peterson L, Ai T, Asatryan B, Bronicki L, Brown E, Celeghin R, Edwards M, Fan J, Ingles J, et al. Evidence-Based Assessment of Genes in Dilated Cardiomyopathy. Circulation. 2021;144:7–19. doi: 10.1161/circulationaha.120.053033

10. Chung H, Kim Y, Park CH, Kim JY, Min PK, Yoon YW, Kim TH, Lee BK, Hong BK, Rim SJ, et al. Genetic relevance and determinants of mitral leaflet size in hypertrophic cardiomyopathy. Cardiovasc Ultrasound. 2019;17:21. doi: 10.1186/s12947-019-0171-1

11. Walsh R, Thomson KL, Ware JS, Funke BH, Woodley J, McGuire KJ, Mazzarotto F, Blair E, Seller A, Taylor JC, et al. Reassessment of Mendelian gene pathogenicity using 7,855 cardiomyopathy cases and 60,706 reference samples. Genet Med. 2017;19:192–203. doi: 10.1038/gim.2016.90

12. Andrews S. FastQC: A quality control tool for high throughput sequence data. In; 2010.

13. Bolger AM, Lohse M, Usadel B. Trimmomatic: a flexible trimmer for Illumina sequence data. Bioinformatics. 2014;30:2114–2120. doi: 10.1093/bioinformatics/btu170

14. Li H. Aligning sequence reads, clone sequences and assembly contigs with BWA-MEM. arXiv preprint arXiv:13033997. 2013.

15. Picard Tools. https://broadinstitute.github.io/picard/.

16. DePristo MA, Banks E, Poplin R, Garimella KV, Maguire JR, Hartl C, Philippakis AA, del Angel G, Rivas MA, Hanna M, et al. A framework for variation discovery and genotyping using next-generation DNA sequencing data. Nat Genet. 2011;43:491–498. doi: 10.1038/ng.806

17. Koboldt DC, Zhang QY, Larson DE, Shen D, McLellan MD, Lin L, Miller CA, Mardis ER, Ding L, Wilson RK. VarScan 2: Somatic mutation and copy number alteration discovery in cancer by exome sequencing. Genome Res. 2012;22:568–576. doi: 10.1101/gr.129684.111

18. Li H. A statistical framework for SNP calling, mutation discovery, association mapping and population genetical parameter estimation from sequencing data. Bioinformatics. 2011;27:2987–2993. doi: 10.1093/bioinformatics/btr509

19. Plagnol V, Curtis J, Epstein M, Mok KY, Stebbings E, Grigoriadou S, Wood NW, Hambleton S, Burns SO, Thrasher AJ, et al. A robust model for read count data in exome sequencing experiments and implications for copy number variant calling. Bioinformatics. 2012;28:2747–2754. doi: 10.1093/bioinformatics/bts526

20. Wang K, Li M, Hakonarson H. ANNOVAR: functional annotation of genetic variants from high-throughput sequencing data. Nucleic acids research. 2010;38:e164. doi: 10.1093/nar/gkq603

21. Morales A, Kinnamon DD, Jordan E, Platt J, Vatta M, Dorschner MO, Starkey CA, Mead JO, Ai T, Burke W, et al. Variant Interpretation for Dilated Cardiomyopathy: Refinement of the American College of Medical Genetics and Genomics/ClinGen Guidelines for the DCM Precision Medicine Study. Circulation Genomic and precision medicine. 2020;13:e002480. doi: 10.1161/circgen.119.002480

22. Richmond CM, James PA, Pantaleo SJ, Chong B, Lunke S, Tan TY, Macciocca I. Clinical and laboratory reporting impact of ACMG-AMP and modified ClinGen variant classification frameworks in MYH7-related cardiomyopathy. Genet Med. 2021. doi: 10.1038/s41436-021-01107-y

23. Whiffin N, Minikel E, Walsh R, O’Donnell-Luria AH, Karczewski K, Ing AY, Barton PJR, Funke B, Cook SA, MacArthur D, et al. Using high-resolution variant frequencies to empower clinical genome interpretation. Genet Med. 2017;19:1151–1158. doi: 10.1038/gim.2017.26

24. Whiffin N, Walsh R, Govind R, Edwards M, Ahmad M, Zhang X, Tayal U, Buchan R, Midwinter W, Wilk AE, et al. CardioClassifier: disease- and gene-specific computational decision support for clinical genome interpretation. Genet Med. 2018;20:1246–1254. doi: 10.1038/gim.2017.258

25. Ioannidis NM, Rothstein JH, Pejaver V, Middha S, McDonnell SK, Baheti S, Musolf A, Li Q, Holzinger E, Karyadi D, et al. REVEL: An Ensemble Method for Predicting the Pathogenicity of Rare Missense Variants. Am J Hum Genet. 2016;99:877–885. doi: 10.1016/j.ajhg.2016.08.016

26. Pejaver V, Byrne AB, Feng BJ, Pagel KA, Mooney SD, Karchin R, O’Donnell-Luria A, Harrison SM, Tavtigian SV, Greenblatt MS, et al. Calibration of computational tools for missense variant pathogenicity classification and ClinGen recommendations for PP3/BP4 criteria. Am J Hum Genet. 2022;109:2163–2177. doi: 10.1016/j.ajhg.2022.10.013

27. Jordan E, Hershberger RE. Considering complexity in the genetic evaluation of dilated cardiomyopathy. Heart. 2021;107:106–112. doi: 10.1136/heartjnl-2020-316658

28. Protonotarios A, Bariani R, Cappelletto C, Pavlou M, García-García A, Cipriani A, Protonotarios I, Rivas A, Wittenberg R, Graziosi M, et al. Importance of genotype for risk stratification in arrhythmogenic right ventricular cardiomyopathy using the 2019 ARVC risk calculator. Eur Heart J. 2022;43:3053–3067. doi: 10.1093/eurheartj/ehac235

29. van Rijsingen IA, Arbustini E, Elliott PM, Mogensen J, Hermans-van Ast JF, van der Kooi AJ, van Tintelen JP, van den Berg MP, Pilotto A, Pasotti M, et al. Risk factors for malignant ventricular arrhythmias in lamin a/c mutation carriers a European cohort study. J Am Coll Cardiol. 2012;59:493–500. doi: 10.1016/j.jacc.2011.08.078

